# Meta-analysis of the clinical performance of commercial SARS-CoV-2 nucleic acid, antigen and antibody tests up to 22 August 2020

**DOI:** 10.1101/2020.09.16.20195917

**Authors:** I Van Walle, K Leitmeyer, E K Broberg, on behalf of the European COVID-19 microbiological laboratories group

## Abstract

We reviewed the clinical performance of SARS-CoV-2 nucleic acid, viral antigen and antibody tests based on 94739 test results from 157 published studies and 20205 new test results from 12 EU/EEA Member States. Pooling the results and considering only results with 95% confidence interval width ≤5%, we found 4 nucleic acid tests, among which 1 point of care test, and 3 antibody tests with a clinical sensitivity ≥95% for at least one target population (hospitalised, mild or asymptomatic, or unknown). Analogously, 9 nucleic acid tests and 25 antibody tests, among which 12 point of care tests, had a clinical specificity of ≥98%. Three antibody tests achieved both thresholds. Evidence for nucleic acid and antigen point of care tests remains scarce at present, and sensitivity varied substantially. Study heterogeneity was low for 8/14 (57.1%) sensitivity and 68/84 (81.0%) specificity results with confidence interval width ≤5%, and lower for nucleic acid tests than antibody tests. Manufacturer reported clinical performance was significantly higher than independently assessed in 11/32 (34.4%) and 4/34 (11.8%) cases for sensitivity and specificity respectively, indicating a need for improvement in this area. Continuous monitoring of clinical performance within more clearly defined target populations is needed.

## Introduction

Testing is one of the central pillars of public health actions in epidemic and pandemic situations to allow timely identification, contact tracing, and isolation of infectious cases to reduce the spread of infectious diseases. In addition, it allows e.g. estimating disease incidence, disease prevalence, and prevalence and duration of humoral immunity. Reliable testing for severe acute respiratory syndrome coronavirus 2 (SARS-CoV-2) and timely reporting of the data to public health authorities is therefore key for management of the coronavirus disease 2019 (COVID-19) pandemic. This requires appropriate and accurate diagnostic tests, with sufficient availability, for both identifying individuals that are currently infected with SARS-CoV-2 as well as those that have been infected in the past.Timely access to testing, sufficient supply of testing materials, availability of tests and related reagents and consumables as well as high-throughput testing are pivotal in this context.

A large number of commercial tests for SARS-CoV-2 RNA (nucleic acid tests or NATs) or viral antigen detection are currently available, as well as serological tests for SARS-CoV-2 specific antibodies. The various types of tests can be used for different purposes and many of these tests have the CE-IVD certificate that indicates compliance with the European in vitro diagnostics directive (98/79/EC) and can thus be marketed within the EU/EEA countries. In addition, the US Food and Drug Administration has granted emergency use authorisations for use of many commercial tests in the US and the World Health Organisation maintains an emergency use listing of commercial tests.[1,2] It is however important to note that the CE-marking is based on a self-declaration of the test manufacturer, including the claims on performance of the test. Independent information on the clinical performance of these tests in terms of sensitivity and specificity is still limited, and yet this is critical for proper interpretation of results.

For this reason, the European Centre for Disease Prevention and Control (ECDC) launched a continuous call to EU/EEA Member States and the UK on 1 April 2020 to provide any such clinical performance data for sharing with other Member States. These data, provided by 12 Member States, are presented in this paper. In addition, publicly available data were added. Finally, minimal performance criteria for different intended uses were gathered from public sources and aided by a survey conducted among EU/EEA member states and the UK from 20 May to 1 June 2020.

## Methods

### Search strategy and selection criteria

Studies containing potentially usable data on clinical performance of SARS-CoV-2 nucleic acid, antigen and antibody tests were first extracted from systematic reviews on this topic. These reviews were identified through an initial Pubmed (Medline) search for systematic reviews and meta-analysis for ‘COVID-19’ and ‘SARS-CoV-2’, followed by snowballing using the ‘find similar articles’ feature. The selection was then extended with the studies listed in the Foundation for Innovative Diagnostics (FIND) database and the European Commission (EC) COVID-19 In Vitro Diagnostic Devices and Test Methods Database. Both databases attempt to exhaustively identify peer-reviewed as well as grey literature on clinical performance of COVID-19 tests and are continuously updated.[3,4] Results from the latter were further filtered on those with a description indicating that they contain clinical performance results. Results produced by United States Food and Drug Administration were also included.[5] Finally, Pubmed was searched according to the query in supplementary Figure S1.

The resulting studies were subsequently assessed for eligibility. At present there are virtually no clinical performance studies that can be judged as being at low risk of bias and low applicability concerns, and systematic reviews up to this point have not used risk of bias or applicability concerns as exclusion criteria.[6-9] This was not done in this work either. Instead, studies were excluded if they did not contain data on commercial tests, or if one or more of the authors were employed by the developer or manufacturer of the index test, to avoid possible conflicts of interest. Studies with an ineligible design, such as blinded tests, analytical validation only, use of another threshold for positivity than in the instructions for use, comparisons between different specimen types or use of an antibody rather than nucleic acid test as reference test for any type of index test were subsequently excluded as well.

Further exclusions were done at sample level based on the reference test employed. Samples classified as actual negatives, i.e. used for determining specificity, had to either (i) be taken before the COVID-19 outbreak, in practice before 2020, (ii) be taken from an individual without COVID-19 compatible symptoms, or (iii) be taken from an individual with COVID-19 compatible symptoms but who was confirmed with another respiratory illness. Samples classified as actual negatives that were taken during the outbreak and were negative according to a nucleic acid test were therefore excluded. This was done to maximally reduce misclassification as actual negatives due to known issues with sensitivity of nucleic acid tests. Such misclassified samples would artificially lower index test specificity in particular when the index test is more sensitive than the reference test.[10-16] For the same reason the reported sensitivity of nucleic acid or antigen index tests, based on a nucleic acid reference test, was considered to be a positive agreement instead, calculated as part of a head-to-head comparison between the two tests. For antibody index tests on the other hand, a nucleic acid test was considered to be a valid reference test to determine actual positive samples and sensitivity, in accordance with WHO interim guidelines.[17]

Manufacturer reported clinical sensitivity and specificity data were extracted from instructions for use where available, or otherwise from the manufacturer’s website. Sensitivity results derived from contrived samples spiked with purified viral RNA were excluded.

### Original clinical performance data

Primary clinical performance data generated by the COVID-19 microbiological laboratories group’s co-authors were assessed by ECDC according to the same criteria as those of the literature review.

### Statistical analysis

Meta-analysis of the included clinical sensitivity and specificity results was performed per test and per target, i.e. the genomic region for nucleic acid tests and the antibody isotype for antibody tests. Antigen targets were not distinguished further. Antibody test sensitivity results below the threshold days after onset were excluded. Sensitivity and positive agreement results were further stratified by case population: hospitalised cases, mild or asymptomatic cases or unknown. Pooled sensitivity and specificity values were calculated using fixed effects analysis, i.e. separately summing and dividing the number of correct predictions by the total number of samples in the group. Wilson 95% confidence intervals were calculated for pooled results. Study heterogeneity was assessed through the I^2^ statistic, calculated through random effects analysis using R version 4.0.2 and the metafor package.[18] I^2^ values <50.0% were considered low heterogeneity, 50.0-74.9% moderate and ≥75% high heterogeneity.

## Results

### Minimum performance criteria

As of 1 June 2020, minimum performance criteria for tests were publicly available from Belgium, France, the Netherlands and the United Kingdom (supplementary Table S1). All were applicable solely to antibody (Ab) tests. The intended uses included diagnosis of COVID-19, determination of exposure to SARS-CoV-2 and determination of the immune status against SARS-CoV-2. Minimum clinical sensitivity for all of the specified intended uses ranged from 85 to 98%, with a median of 95%. These thresholds applied to samples collected at least >14 days post onset of symptoms (dpo), taking into account the time to seroconversion. Minimum clinical specificity for all of the specified intended uses was 98% in six countries and 98.5% in one. For nucleic acid and antigen confirmatory tests, the draft WHO Target Product Profiles for priority diagnostics to support response to the COVID-19 pandemic state >95% - >98% sensitivity (acceptable/desired) and >99% specificity.[19] General thresholds of >95% sensitivity and >98% specificity were used for further analysis, together with a maximum 95% confidence interval (CI) width of ≤5%. For IgM only results, an upper limit of ≤28 dpo, or the highest dpo category having a lower limit ≤28 dpo, was added as well since IgM antibodies decrease fairly rapidly and such tests are not intended to be used long after exposure.[20]

### Primary clinical performance data

Eight systematic reviews were identified, including one by health technology assessment bodies not listed as a peer-reviewed study, and the primary studies included in the analysis extracted.[6-9,21-24] The full list of studies in the FIND and EC databases was retrieved on 22 August 2020. Pubmed was searched on the same date. From the EC database, 268 out 385 studies were screened out since their description did not indicate that they would contain clinical performance data on commercial tests. Analogously, 1520 out of 1738 studies were screened out from the Pubmed results. From the combined list of 364 studies, 99 had no clinical performance data on commercial tests, 34 were excluded due to a potential conflict of interest and 74 were excluded due to an ineligible design, leaving a total 157 included studies. A complete overview of the study selection is given in Figure 1. After exclusion of antibody test sensitivity results ≤14 dpo and ineligible specificity results, a total of 38202 and 56537 index test results remained for calculation of sensitivity and specificity, respectively. After addition of original, previously unpublished results provided by the authors of this study, this increased to 48310 and 66634 index test results, respectively, for 201 tests (Supplementary Tables S2-S4).

**Figure 1.**
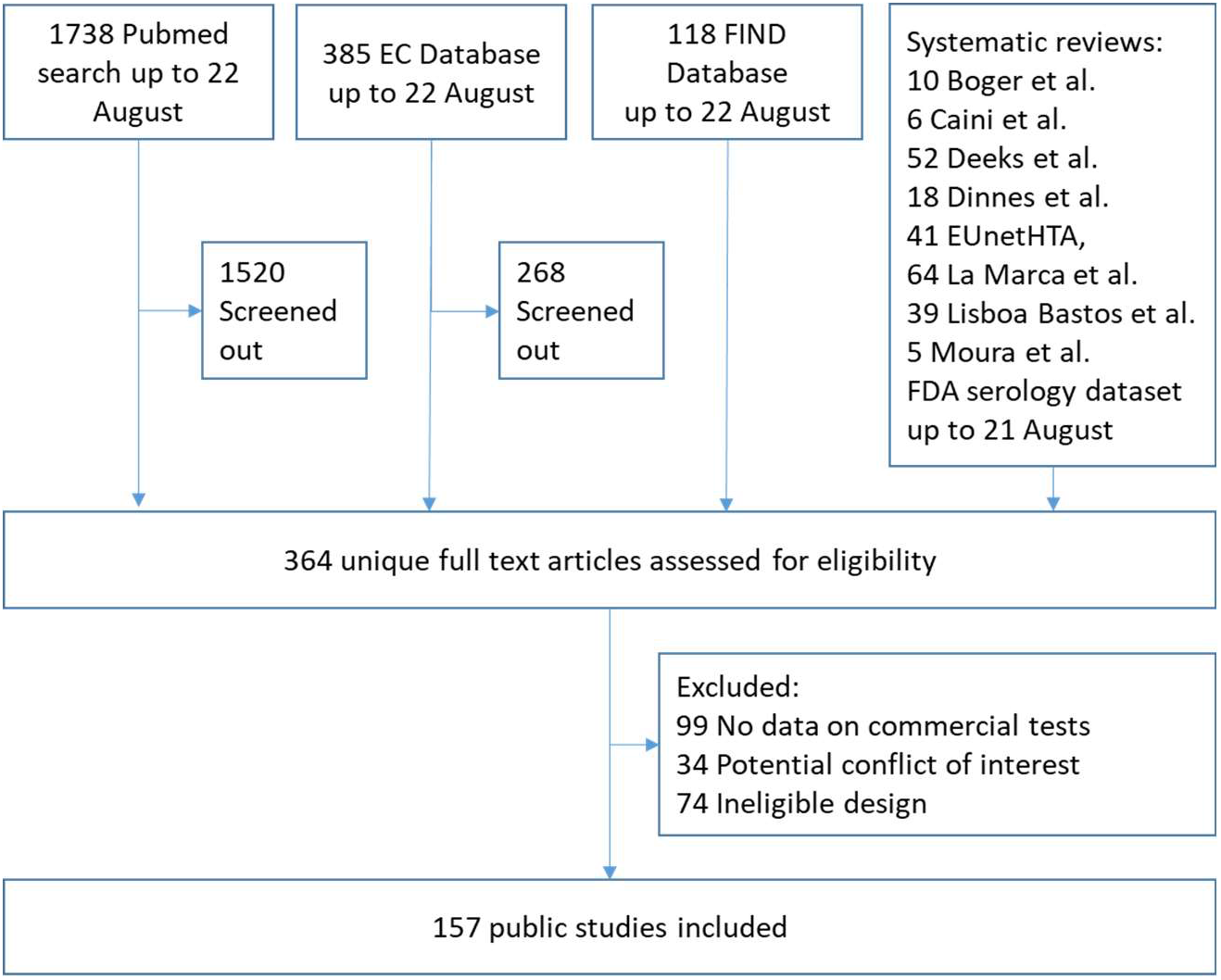
Selection of public studies.

### Meta-analysis

Pooled estimates for clinical sensitivity and specificity per test, target and, for sensitivity, case population were made as described in methods. For antibody tests, results were restricted to those estimates that had a 95% CI width of ≤5% and were derived from at least two studies, to be able to assess study heterogeneity. Based on the minimum performance criteria analysis, results above ≥95% sensitivity and/or ≥98% specificity for a particular population are highlighted (Table 1). Among these results, there were two CLIAs, one ELISA and no LFIAs/POCs, that had ≥95% sensitivity.Analogously, there were nine CLIAs, four ELISAs and twelve LFIAs/POCs, that had ≥98% specificity, among which the three with ≥95% sensitivity. Study heterogeneity was low for 4/10 (40.0%) sensitivity and 53/69 (76.8%) specificity results with CI width of ≤5%. There were few sensitivity results for IgG for mild or asymptomatic cases, IgA and total antibody, none of which had a CI width of ≤5% (Table 1). Compared to the same test used for hospitalised cases, drops in sensitivity were observed of 7.4%, 11.0%, 13.1% and 19.2% for IgG, 28.8% for IgA and -6.0% for total antibody. The latter negative value is likely due to low number of samples for both populations.

**Table 1:**
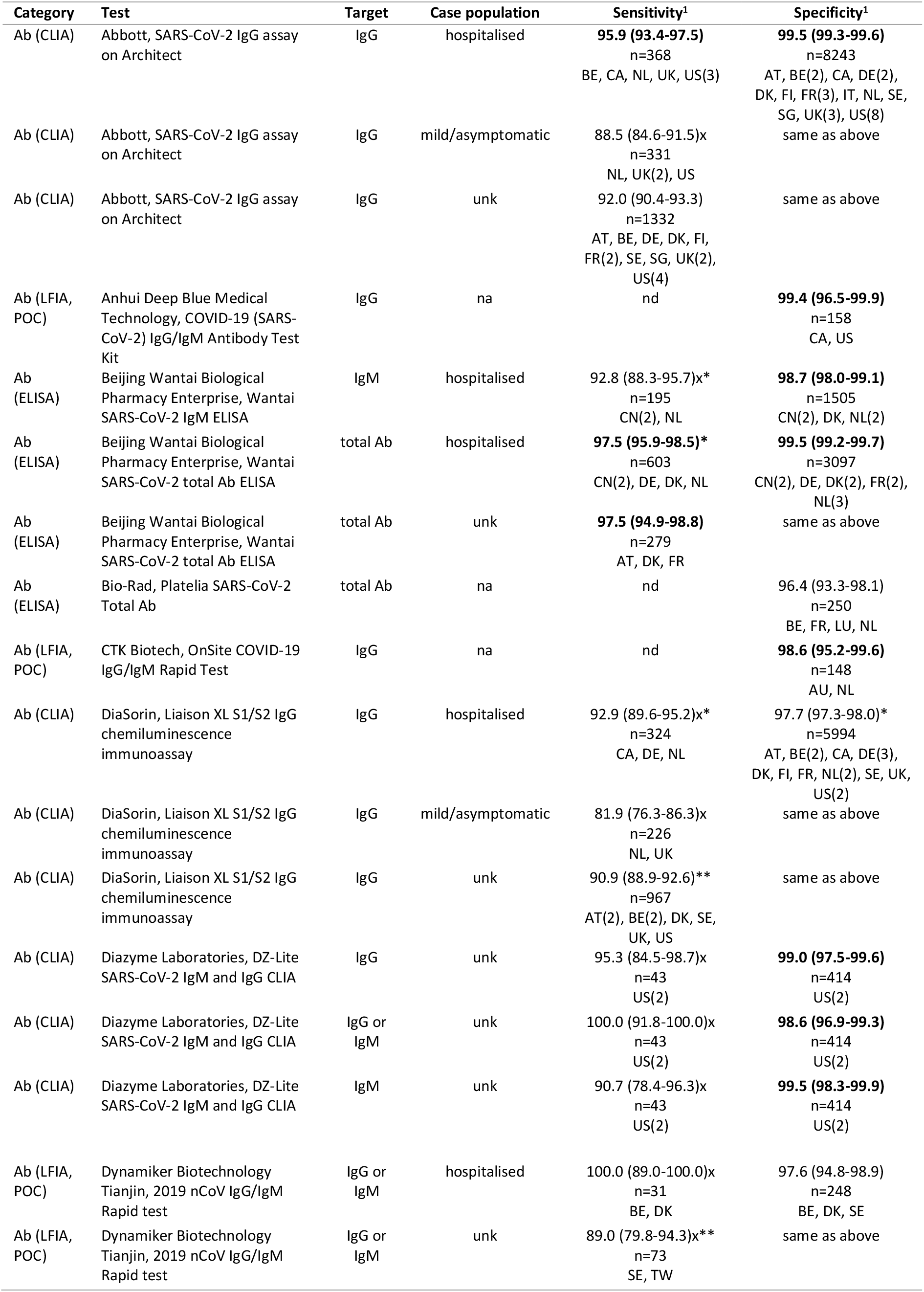
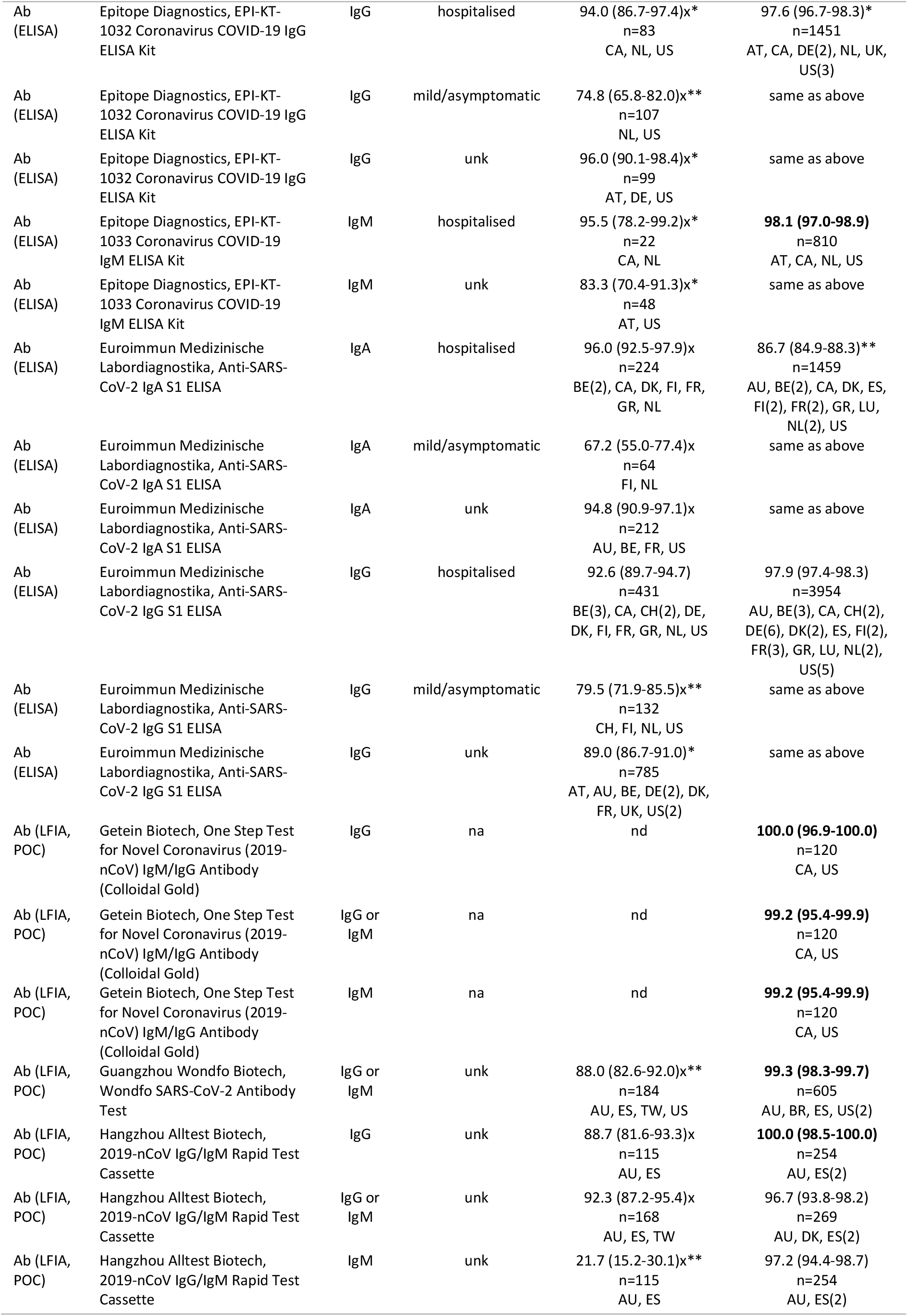
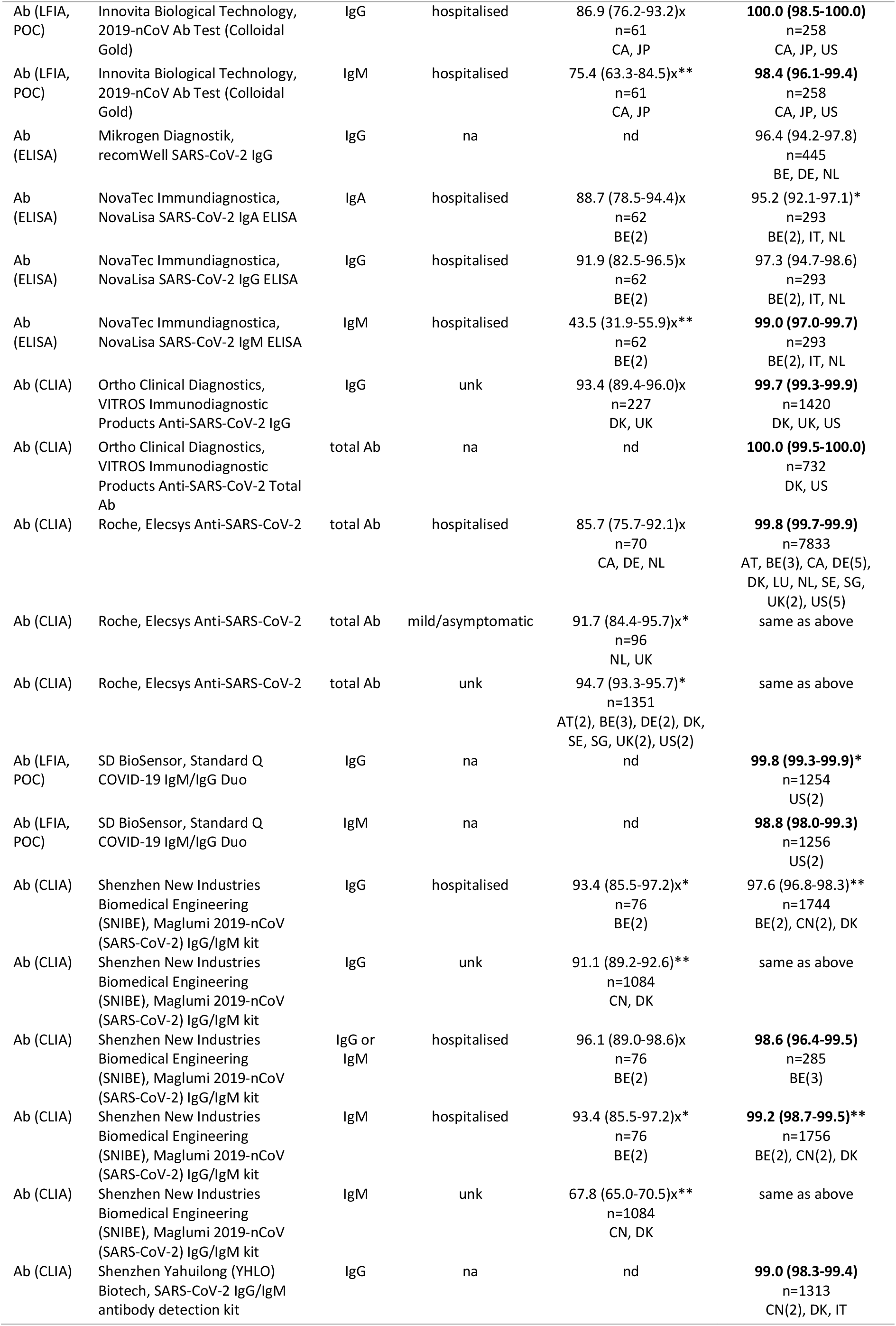
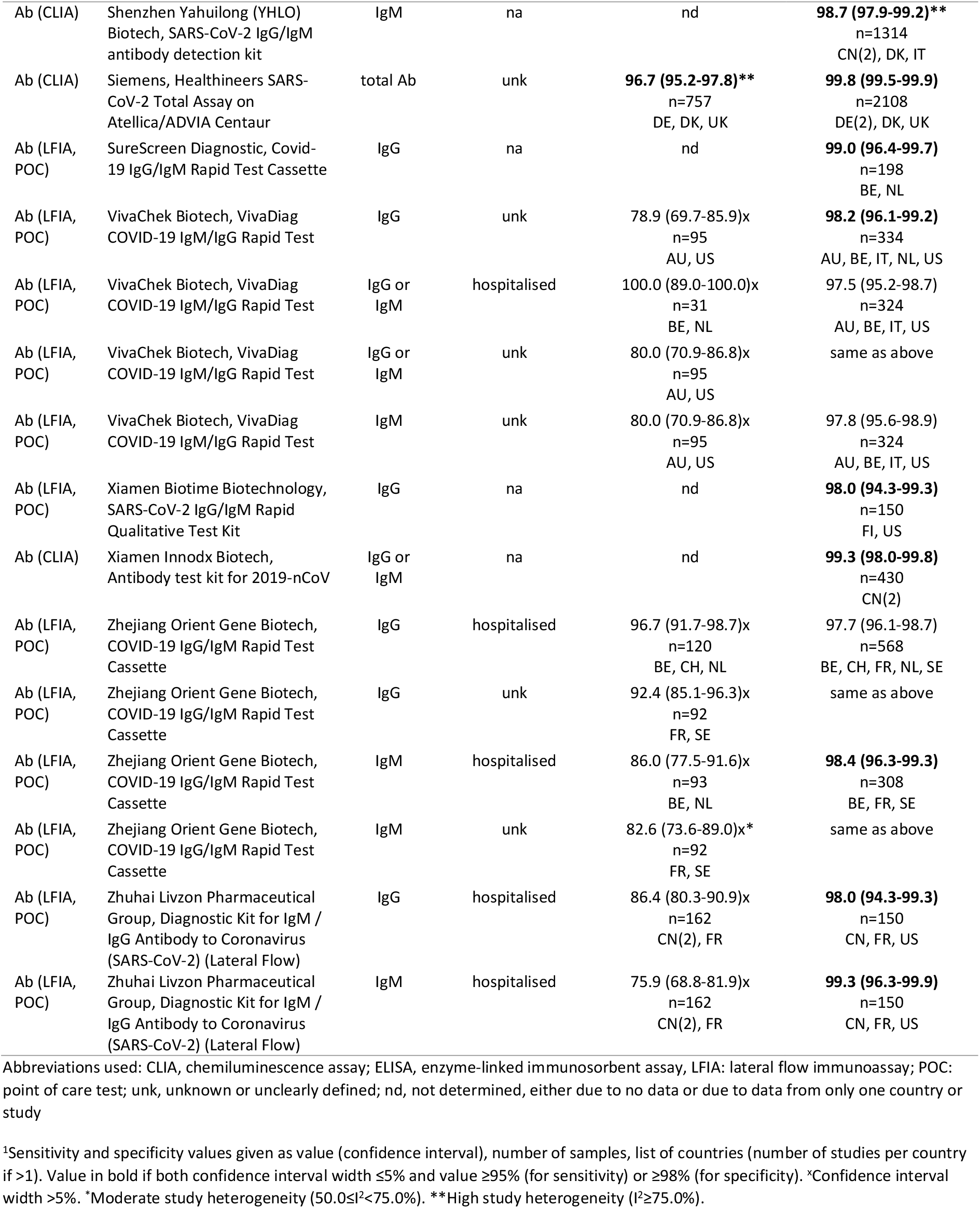
Pooled sensitivity and specificity results for antibody tests with confidence interval width ≤5% for either or both and based on ≥2 studies. Only samples taken >14 days post onset of symptoms are included, and ≤28 days post onset for IgM only as target. Rows are sorted alphabetically by test, target and case population.

For nucleic acid tests that were not point of care (POC) tests, results were restricted as for antibody tests (Table 2). Three tests had ≥95% positive agreement with a CI width of ≤5%, and nine had ≥98% specificity. Study heterogeneity was low for all 4 sensitivity and all 15 specificity results with CI width of ≤5%. Limited data were available for five POC antigen tests and five POC nucleic acid tests (Table 3). Large variability in positive agreement was observed. The best performing nucleic acid POC achieved a positive agreement of 98.9% (97.3-99.6%), i.e. with a CI width of ≤5%. Virtually no data were available on specificity, but no false positives were observed among them for either antigen or nucleic acid POCs.

**Table 2:** Pooled positive agreement and specificity results for nucleic acid tests with confidence interval width ≤5% for either or both and based on ≥2 studies. Rows are sorted alphabetically by test, target and case population.

| Category | Test | Target | Case population | Positive agreement<sup>1</sup> | Specificity<sup>1</sup> |
| --- | --- | --- | --- | --- | --- |
| PCR | Altona Diagnostics, RealStar SARS-CoV-2 RT-PCR Kit 1.0 | E | unk | 88.1 (80.4-93.1)x<br>n=101<br>CH, FR, NL, US | **100.0 (96.7-100.0)**<br>n=112<br>CH, NL |
| PCR | Altona Diagnostics, RealStar SARS-CoV-2 RT-PCR Kit 1.0 | S | unk | 87.1 (79.2-92.3)x<br>n=101<br>CH, FR, NL, US | **100.0 (96.7-100.0)**<br>n=112<br>CH, NL |
| PCR | Altona Diagnostics, RealStar SARS-CoV-2 RT-PCR Kit 1.0 | S or E | unk | 81.6 (75.8-86.3)x*<br>n=207<br>FR(3), NL | **100.0 (98.4-100.0)**<br>n=237<br>FR, NL, UK |
| PCR | AusDiagnostics, Coronavirus Typing Assay | ORF1ab | na | nd | **100.0 (98.5-100.0)**<br>n=254<br>AU, UK |
| PCR | BGI, Real-time fluorescent RT-PCR kit for detecting 2019 nCoV | ORF1ab | unk | 93.8 (88.7-96.7)x<br>n=146<br>CH, JP, NL, PL | **99.1 (95.1-99.8)**<br>n=112<br>CH, NL |
| PCR | CerTest Biotec, VIASURE SARS-CoV-2 Real Time PCR Detection Kit | N | unk | 96.8 (89.1-99.1)x*<br>n=63<br>CH, NL | **100.0 (96.7-100.0)**<br>n=112<br>CH, NL |
| PCR | CerTest Biotec, VIASURE SARS-CoV-2 Real Time PCR Detection Kit | ORF1ab | unk | 93.7 (84.8-97.5)x**<br>n=63<br>CH, NL | **100.0 (96.7-100.0)**<br>n=112<br>CH, NL |
| PCR | CerTest Biotec, VIASURE SARS-CoV-2 Real Time PCR Detection Kit | ORF1ab or N | na | nd | **100.0 (98.2-100.0)**<br>n=207<br>NL, UK |
| PCR | DiaSorin, Simplexa COVID-19 Direct RT-PCR Kit | ORF1ab or S | unk | **97.8 (94.4-99.1)**<br>n=180<br>US(3) | nd |
| PCR | Hologic, SARS-CoV-2 Assay (Panther Fusion System) | ORF1ab | unk | **98.3 (96.8-99.1)**<br>n=525<br>FR, US(6) | nd |
| PCR | KH Medical, RADI COVID-19 Detection Kit and RADI COVID-19 Triple Detection Kit | RdRP | unk | 96.8 (89.1-99.1)x*<br>n=63<br>CH, NL | **100.0 (96.7-100.0)**<br>n=112<br>CH, NL |
| PCR | KH Medical, RADI COVID-19 Detection Kit and RADI COVID-19 Triple Detection Kit | S | unk | 98.4 (91.5-99.7)x<br>n=63<br>CH, NL | **100.0 (96.7-100.0)**<br>n=112<br>CH, NL |
| PCR | Primerdesign, genesig Real-Time PCR CoVID-19 kit | RdRP | unk | 95.3 (89.4-98.0)x*<br>n=106<br>CH, NL, PL | **100.0 (98.8-100.0)**<br>n=307<br>CH, NL, UK |
| PCR | R-Biopharm, Ridagene SARS-CoV2 | E | unk | 100.0 (94.3-100.0)x<br>n=63<br>CH, NL | **100.0 (96.7-100.0)**<br>n=112<br>CH, NL |
| PCR | Roche, COBAS SARS-CoV-2 test | ORF1ab or E | unk | **98.8 (97.9-99.3)**<br>n=1125<br>AT, CH, DE, FR, SI, US(5) | 100.0 (90.8-100.0)x<br>n=38<br>CH, FR |
| PCR | Seegene, Allplex 2019-nCoV assay | E | unk | 85.0 (75.6-91.2)x**<br>n=80<br>CH, FR, NL | **100.0 (96.7-100.0)**<br>n=112<br>CH, NL |
| PCR | Seegene, Allplex 2019-nCoV assay | RdRP | unk | 91.3 (83.0-95.7)x*<br>n=80<br>CH, FR, NL | **100.0 (96.7-100.0)**<br>n=112<br>CH, NL |
| PCR | Tibmolbiol, SARS-CoV (COVID19) E-gene | E | unk | 100.0 (94.4-100.0)x<br>n=65<br>CH, UK | **100.0 (98.5-100.0)**<br>n=250<br>CH, UK |

Abbreviations used: PCR, polymerase chain reaction; unk, unknown or unclearly defined; nd, not determined, either due to no data or due to data from only one country or study
<sup>1</sup>Positive agreement and specificity values given as value (confidence interval), number of samples, list of countries (number of studies per country if >1). Value in bold if both confidence interval width ≤5% and value ≥95% (for positive agreement) or ≥98% (for specificity). <sup>\*</sup>Confidence interval width >5%. <sup>\*</sup>Moderate study heterogeneity (50.0≤I<sup>2</sup><75.0%). <sup>\*\*</sup>High study heterogeneity (I<sup>2</sup>≥75.0%).

**Table 3:**
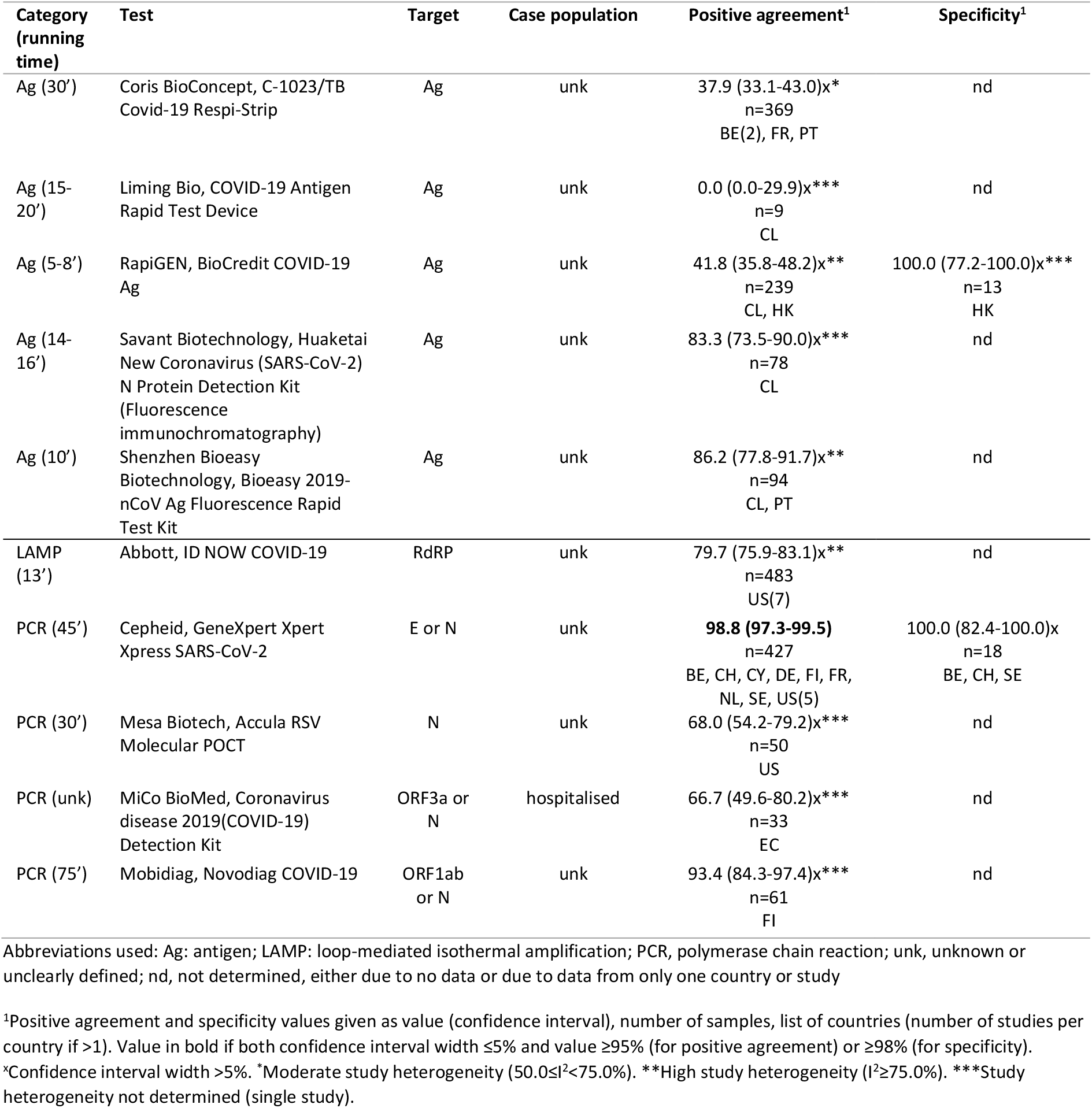
Pooled positive agreement and specificity results for point of care antigen and nucleic acid tests. Rows are sorted alphabetically by category, test, target and case population.

The correlation between independently assessed clinical performance results and manufacturer reported results is shown in Figure 2. Only independently assessed results with CI width of ≤5% are included. A total of 11/32 (34.4%) of sensitivity and 4/33 (12.1%) specificity results were significantly different (p<0.05).

**Figure 2.**
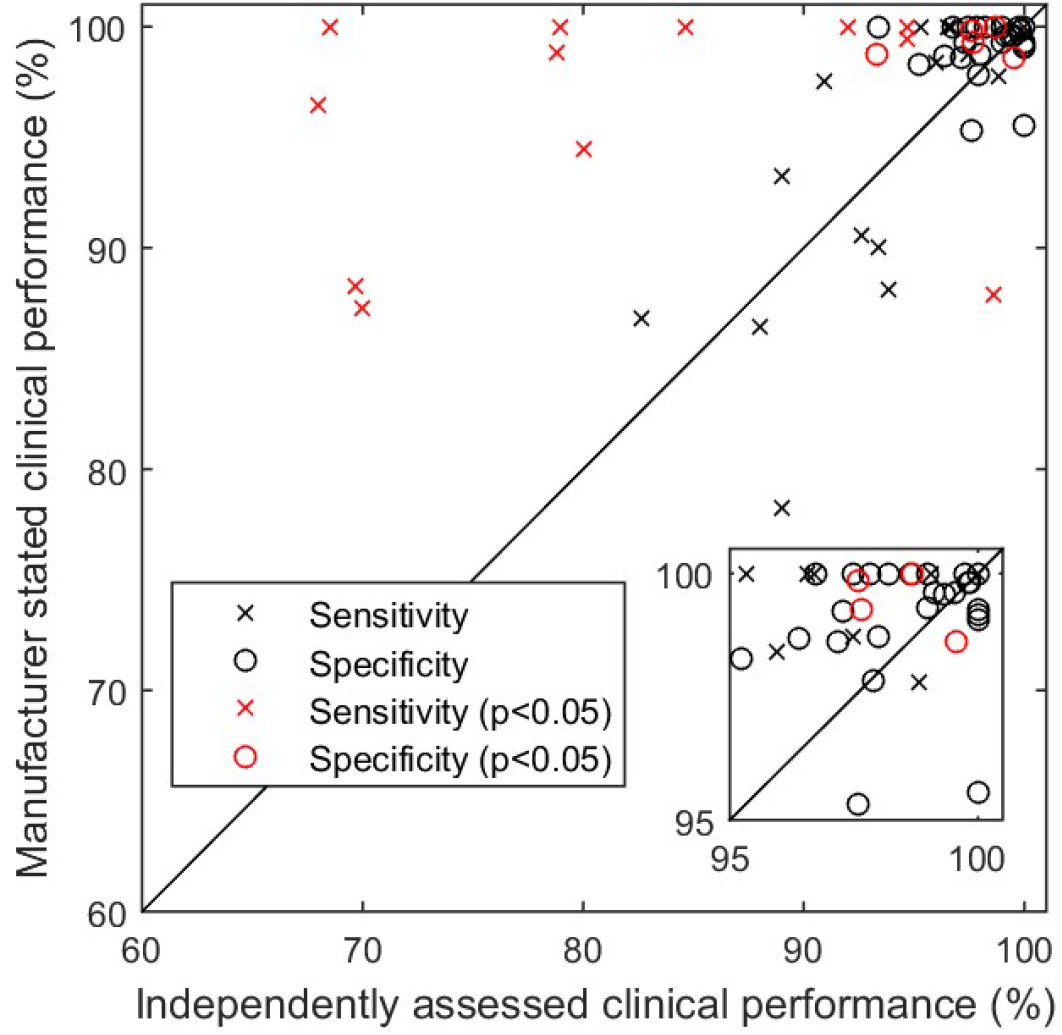
Independently assessed versus manufacturer reported clinical sensitivity and specificity per test, with significantly different (p<0.05) results highlighted. Independently assessed results limited to those with 95% confidence interval width of ≤5%. The inset expands the 95-100% region.

## Discussion

This review represents, to our knowledge, the most complete independent overview so far of clinical performance of commercially available COVID-19 tests. A substantial amount of previously unpublished data from European countries are included as well. To date, there are numerous commercial tests for which sufficient performance data are available to allow calculation of clinical sensitivity or positive agreement, and specificity with narrow confidence interval ranges.Reassuringly, the clinical performances of several nucleic acid and antibody tests exceeded the minimum performance criteria. As time progresses, the list of tests with sufficient available performance data is expected to increase.

At the same time, the available evidence for point of care nucleic acid and antigen tests remains scarce, even though these tests can have substantial practical advantages for e.g. screening. We therefore recommend more emphasis on the validation of these tests, including as part of a testing algorithm, whereby the sensitivity and specificity of taking two tests with a number of days in between is assessed, and which can e.g. be useful to reduce the duration of a quarantine period.

The comparison between the independently assessed clinical performance data and manufacturer reported clinical performance revealed that in particular sensitivity is frequently (40.7% of the cases in this study) significantly overestimated by the manufacturer. At a minimum, this emphasises that such independent assessments are clearly necessary. In the longer term, an explicit and proactive regulatory mechanism in Europe to compare available independently generated evidence on these tests with manufacturer reported values, coupled with appropriate regulatory action, could be useful. This could also be rewarding towards those manufacturers that do provide robust estimates of their product’s performance.

Limitations of this paper include that most of the included studies have a substantial risk of bias in the sample selection, especially for the sensitivity panel, as established also in the assessments performed in the systematic reviews that were used as a source. Results were mainly based on hospitalised cases or poorly defined populations, whereas the population of interest consists often of symptomatic cases in general or even asymptomatic cases, and differences in performance may exist depending on disease severity. While this review addresses a pressing need for actionable clinical performance data, ideally, the clinical performance should be assessed through prospective studies or clinical trials with a guaranteed unbiased sample selection for a clearly defined target population and intended use of the test. Given the difficulty of assessing and extracting the data from individual studies in a coherent way, we recommend that the Standard for Reporting of Diagnostic Accuracy Studies (STARD) should also be followed when publishing the results.[25]

In this context, the selection of the reference test is particularly important with respect to reference negative samples. As described in some of the assessed studies, it should be avoided that index test results are considered as false positives while the samples are from actual cases and for this reason we excluded nucleic acid negative samples from suspected COVID-19 patients altogether. We expect therefore little bias in the specificity results, except potentially from under or overrepresentation of confounders. This is especially relevant for seroprevalence studies, where, in the current low-prevalence situation, in particular the specificity of the test needs to be well-defined and high. On the other hand, sensitivity results using a nucleic acid test as reference should be interpreted with caution, since the positive samples may exclude some actual cases.

Possibilities to improve the reference test can include testing - potentially only the false positives - with a second reference nucleic acid test preferably targeting different genes, testing more than one sample from the same patient including for antibodies at a later time point, testing samples from both upper and lower respiratory tracts, and sequencing the sample. The handling of intermediate index test results is an issue that needs to be described in studies, and in general these should be considered as positive results rather than either negatives or being excluded from the validation, since they would normally require further follow-up to confirm the positivity of the sample.

For the above reasons, the authors and organisations contributing to this study in no way recommend the use of the listed commercial tests over other not listed commercial or in-house tests. Finally, it should be kept in mind that this study is a snapshot in time and that the clinical performance of tests may change over time as the virus population evolves. We therefore recommend a continuous monitoring of clinical performance both in Europe and globally, which is key for reliable monitoring of the pandemic and which will also support vaccine and antiviral development. These results should be shared in a timely manner publically

## Data Availability

All data are either publicly available or included in the supplementary material

## Acknowledgements

We would like to acknowledge the technicians in the European microbiology laboratories who work hard to support the control of COVID-19 and supported the validations described in this manuscript.

We would like to acknowledge the companies that made some of the kits available for evaluation to some of the laboratories.

## European COVID-19 microbiological laboratories group

Marjan Van Esbroeck (Institute of Tropical Medicine, Antwerpen, Belgium), Pieter Vermeersch (Clinical Department of Laboratory Medicine and National Reference Center for Respiratory Pathogens, University Hospitals Leuven, Leuven, Belgium), Kurt Beuselinck (Clinical Department of Laboratory Medicine and National Reference Center for Respiratory Pathogens, University Hospitals Leuven, Leuven, Belgium), Christos Karagiannis (Nicosia General Hospital, Cyprus), Merit Melin (Department of Health Protection, Expert Microbiology Unit, Finnish Institute for Health and Welfare (THL), Helsinki, Finland), Nina Ekström (Department of Health Protection, Expert Microbiology Unit, Finnish Institute for Health and Welfare (THL), Helsinki, Finland), Iris Erlund (Department of Government Services, Forensic Toxicology Unit, Finnish Institute for Health and Welfare (THL), Helsinki, Finland), Terhi Vihervaara (Department of Government Services, Forensic Toxicology Unit, Finnish Institute for Health and Welfare (THL), Helsinki, Finland), Vanessa Escuret (Laboratoire de Virologie des HCL, Institut des Agents Infectieux, CNR des virus à transmission respiratoire (dont la grippe), Groupement Hospitalier Nord, Lyon, France), Emilie Frobert (Laboratoire de Virologie des HCL, Institut des Agents Infectieux, CNR des virus à transmission respiratoire (dont la grippe), Groupement Hospitalier Nord, Lyon, France), Alexandre Gaymard (Laboratoire de Virologie des HCL, Institut des Agents Infectieux, CNR des virus à transmission respiratoire (dont la grippe), Groupement Hospitalier Nord, Lyon, France), Andreas Mentis (Hellenic Pasteur Institute, Athens, Greece), Stavroula Lampropoulou (Hellenic Pasteur Institute, Athens, Greece), Ivan-Christian Kurolt (Research unit, University Hospital for Infectious Diseases “Dr. Fran Mihaljević”, Zagreb, Croatia), Tamir Abdelrahman (Department of Microbiology, Laboratoire national de santé, Luxembourg), Trung Nguyen (Department of Microbiology, Laboratoire national de santé, Luxembourg), Guillaume Fournier (Department of Microbiology, Laboratoire national de santé, Luxembourg), Chantal B.E.M. Reusken (Centre for Infectious Disease Control, National Institute for Public Health and the Environment, The Netherlands), Maaike J.C. van den Beld (Centre for Infectious Disease Control, National Institute for Public Health and the Environment, The Netherlands), Janette Rahamat-Langendoen (Department of Medical Microbiology, Radboud University Medical Center, Nijmegen), Marjolijn C.A. Wegdam-Blans (Department of Medical Microbiology, PAMM, Veldhoven, The Netherlands), Jeroen H. T. Tjhie (Department of Medical Microbiology, PAMM, Veldhoven, The Netherlands), Peter Croughs (Department of Medical Microbiology and Infectious Diseases, Erasmus Medical Center, Rotterdam, The Netherlands), Corine H. GeurtsvanKessel (Department of Virology, Erasmus Medical Center, Rotterdam, The Netherlands), Johan Reimerink (Centre for Infectious Diseases Research, Diagnostics and Laboratory Surveillance, Centre for Infectious Disease Control, National Institute for Public Health and the Environment, The Netherlands), David S.Y. Ong ([a] Department of Medical Microbiology and Infection Control, Franciscus Gasthuis & Vlietland, Rotterdam, The Netherlands, [b] Department of Epidemiology, Julius Center for Health Sciences and Primary Care, University Medical Center Utrecht, Utrecht, The Netherlands), Hans G.M. Koeleman (Department of Medical Microbiology and Infection Control, Franciscus Gasthuis & Vlietland, Rotterdam, The Netherlands), Hannke Berkhout (Canisius-Wilhelmina hospital, Nijmegen, The Netherlands), Christel F.M. van der Donk (Canisius-Wilhelmina hospital, Nijmegen, The Netherlands), Menno D. de Jong (Department of Medical Microbiology & Infection prevention, Amsterdam University Medical Centers, The Netherlands), Rens Zonneveld (Department of Medical Microbiology, Amsterdam University Medical Center, Amsterdam, The Netherlands), Suzanne Jurriaans (Department of Medical Microbiology, Amsterdam University Medical Center, Amsterdam, The Netherlands), Nathalie Van Burgel (Hagaziekenhuis, The Hague, The Netherlands), Bas B. Wintermans (Department of Medical Microbiology and Immunology, Admiraal de Ruyter Hospital, Vlissingen, The Netherlands), Ger T. Rijkers ([a] Department of Medical Microbiology and Immunology, Admiraal de Ruyter Hospital, Goes, The Netherlands, [b] Elisabeth-Tweesteden Hospital, Tilburg, The Netherlands), Jean-Luc Murk (Elisabeth-Tweesteden Hospital, Tilburg, The Netherlands), Khoa T.D. Thai ([a] Unit of Medical Microbiology, Star-shl Medical Diagnostic Center, Rotterdam, The Netherlands, [b] Department of Medical Microbiology and Infectious Diseases, Erasmus Medical Center, Rotterdam, The Netherlands), Melanie J de Graaf ([a] Department of Medical Microbiology, University Medical Centre, Utrecht, The Netherlands, [b] Saltro Diagnostic Centre, Utrecht, The Netherlands), Annemarie van’t Veen ([a] Department of Medical Microbiology, University Medical Centre, Utrecht, The Netherlands, [b] Saltro Diagnostic Centre, Utrecht, The Netherlands), Cornelis P. Timmerman (Central Bacteriology and Serology Laboratory, Tergooi Hospital, Hilversum, The Netherlands), Annette van Corteveen-Splinter (Central Bacteriology and Serology Laboratory, Tergooi Hospital, Hilversum, The Netherlands), Felix Geeraedts (Laboratory for Medical Microbiology and Public Health, Hengelo, The Netherlands), Adrian Klak (Laboratory for Medical Microbiology and Public Health, Hengelo, The Netherlands), Maria M. Konstantinovski (Reinier Haga Medical Diagnostic Centre, Delft, The Nederlands), Manou R. Batstra (Reinier Haga Medical Diagnostic Centre, Delft, The Nederlands), K. A. Heemstra (Alrijne Zorggroep, Leiderdorp, The Netherlands), Jos J. Kerremans (Alrijne Zorggroep, Leiderdorp, The Netherlands), Inge H. M. van Loo ([a] Department of Medical Microbiology, Maastricht University Medical Center, The Netherlands, [b] Care and Public Health Research Institute, Maastricht University), Paul H. M. Savelkoul ([a] Department of Medical Microbiology, Maastricht University Medical Center, The Netherlands, [b] Care and Public Health Research Institute, Maastricht University), Johan Kissing (Department of Medical Microbiology and Infection prevention, Gelre Hospitals, Apeldoorn, The Netherlands), Paul Martijn den Reijer (Department of Medical Microbiology and Infection prevention, Gelre Hospitals, Apeldoorn, The Netherlands), Anne Russcher (Department of Medical Microbiology, Medical Meander Center, Amersfoort, The Netherlands), Moniek Heusinkveld (Department of Medical Microbiology, Hospital Gelderse Vallei, Ede, The Netherlands), Ellen van Lochem (Department of Medical Microbiology and Immunology, Hospital Rijnstate, The Netherlands), Steven F. T. Thijsen (Medical Microbiology and Immunology, Diakonessen Hospital, Utrecht, The Netherlands), Michiel Heron (Medical Microbiology and Immunology, Diakonessen Hospital, Utrecht, The Netherlands), Susanne P. Stoof (Department of Medical Microbiology, Comicro, Hoorn, The Netherlands), Sim van Gyseghem (Department of Medical Microbiology, Comicro, Hoorn, The Netherlands), Sylvia B. Debast (Laboratory of Clinical Microbiology and Infectious Diseases, Isala Hospital, Zwolle, The Netherlands), Claudy Oliveira dos Santos (Laboratory of Clinical Microbiology and Infectious Diseases, Isala Hospital, Zwolle, The Netherlands), Bjorn L. Herpers (Regional Public Health Laboratory Kennemerland, The Netherlands), Theo Mank (Regional Public Health Laboratory Kennemerland, The Netherlands), Kin Ki Jim ([a] Department of Medical Microbiology and Infection Control, Jeroen Bosch Hospital, ‘s-Hertogenbosch, The Netherlands, [b] Department of Medical Microbiology and Infection Prevention, Amsterdam University Medical Centers, Amsterdam institute for Infection and Immunity, Amsterdam, The Netherlands), Peter C. Wever (Department of Medical Microbiology and Infection Control, Jeroen Bosch Hospital, ‘s-Hertogenbosch, The Netherlands), Jutte J.C. de Vries (Department of Medical Microbiology, Leiden University Medical Center, Leiden, The Netherlands), Martine Hoogewerf (Department of Medical Microbiology, Northwest Hospital Group, Alkmaar, The Netherlands), Deborah J. Kaersenhout (Atalmedial Medical Microbiology Laboratory, Amsterdam, the Netherlands), Annette M. Stemerding (Deventer Ziekenhuis, Deventer, the Netherlands), Babette C. van Hees (Deventer Ziekenhuis, Deventer, the Netherlands), Vishal Hira (Department of Medical Microbiology and Infection Prevention, Groene Hart Ziekenhuis, Gouda, the Netherlands), Anne E. Bos (Department of Medical Microbiology and Infection Prevention, Groene Hart Ziekenhuis, Gouda, the Netherlands), Leontine Mulder (Clinical Laboratory, Medlon B.V., Enschede, The Netherlands), Michiel van Rijn (Medical Laboratory, Ikazia Hospital, Rotterdam, The Netherlands), Aleksander Michalski (Epidemiological Response Centre of The Polish Armed Forces, Warsaw, Poland), Marta Pakiela (Voivodeship Sanitary Epidemiological Station, Warsaw, Poland), Anna Siewierska-Puchlerska (Voivodeship Sanitary Epidemiological Station, Warsaw, Poland), Jarosław Paciorek (Voivodeship Sanitary Epidemiological Station, Warsaw, Poland), Ewa Gajda (Epidemiological Response Centre of The Polish Armed Forces, Warsaw, Poland), Katarzyna Pancer (Department of Virology, BSL3 Laboratory, COVID-19 NIPH-NIH team, National Institute of Public Health-National Institute of Hygiene, Warsaw, Poland), Agnieszka Kołakowska-Kulesza (Department of Virology, COVID-19 NIPH-NIH team, National Institute of Public Health-National Institute of Hygiene, Warsaw, Poland), Magdalena Rzeczkowska (Department of Bacteriology and Biocontamination Control, COVID-19 NIPH-NIH team, National Institute of Public Health-National Institute of Hygiene, Warsaw, Poland), Raquel Guiomar (Instituto Nacional de Saúde Dr. Ricardo Jorge, I.P., Portugal.), Líbia Zé-Zé (Instituto Nacional de Saúde Dr. Ricardo Jorge, I.P., Portugal.), Inês Costa (Instituto Nacional de Saúde Dr. Ricardo Jorge, I.P., Portugal.), Johan Brynedal Öckinger (Department of Virology, Clinical Microbiology, Karolinska University Laboratory, Karolinska University Hospital, Stockholm, Sweden), Berit Hammas (Department of Virology, Clinical Microbiology, Karolinska University Laboratory, Karolinska University Hospital, Stockholm, Sweden), Katarina Prosenc (National Laboratory for Health, Environment and Food Slovenia, Laboratory for Public Health Virology), Nataša Berginc (National Laboratory for Health, Environment and Food Slovenia, Laboratory for Public Health Virology)

## Authors’ contributions

Ivo Van Walle: conceptualisation, methodology, data curation, formal analysis, writing-review, editing. Katrin Leitmeyer: conceptualisation, methodology, data curation, writing-review, editing. Eeva K. Broberg: conceptualisation, methodology, data curation, writing-review, editing. European COVID-19 microbiological laboratories group: conceptualization, investigation, data curation, methodology, writing-review.

---

**SUPPLEMENTARY FIGURE S1**: Pubmed search string

**SUPPLEMENTARY TABLE S1**: Minimum performance criteria proposed by different institutes. The data are summarised and translated when needed (only the original text is valid).

**SUPPLEMENTARY TABLE S2**: Studies included in the meta-analysis.

**SUPPLEMENTARY TABLE S3**: Clinical performance results.

**SUPPLEMENTARY TABLE S4**: Forest plots.

